# Trends in Gaps of Care for Congenital Heart Disease Patients: Implications for Social Determinants of Health and Child Opportunity Index

**DOI:** 10.1101/2024.02.06.24302427

**Authors:** Abbas H. Zaidi, Adam Alberts, Devyani Chowdhury, Claude Beaty, Benjamin Brewer, Ming Hui Chen, Sarah D. de Ferranti

## Abstract

**Background:** Lifelong continuity of care is imperative for patients with congenital heart disease (CHD); unfortunately, gaps in care (GIC) are common.

**Methods:** All patients aged 0-34 years followed at a pediatric subspecialty hospital (primary location Delaware; satellites covering Pennsylvania, New Jersey, Maryland) with CHD who underwent surgery between January 2003 and May 2020 were included. Patients were categorized as simple, moderate, and complex CHD based on 2018 American Heart Association and American College of Cardiology guidelines. Social determinants of health factors such as age, race, ethnicity, sex, language, insurance status, and Child Opportunity Index based on home address zip code were analyzed.

**Results:** Of 2012 CHD patients, a GIC of ≥3 years was identified in 56% (n=1119). The proportion of patients with GIC per year increased for all patients. Multivariable longitudinal models with all CHD patients showed that GIC are increasing for patients who are ≥10.5 years old, have simple CHD, live out of state, live farther from a site of care (hospital or satellite clinics), receive public insurance and/or have less protection with additional insurance plans, and reside in low Child Opportunity Index neighborhoods. A separate model for only moderate/complex CHD patients showed similar findings. Neither longitudinal model showed race/ethnicity as significant for increasing GIC trends.

**Conclusions:** GIC have continued to increase with an aging CHD population with social determinants of health factors specifically related to insurance, access, and neighborhood opportunity. Attenuating GIC necessitates standardized practices while simultaneously addressing the impact of SDOH on all CHD patients.

**Clinical Perspective:** *What is new?:*

- This study is the first to examine gaps in care (GIC) trends over time in patients with repaired congenital heart disease (CHD) and found that almost half had GIC.
- Over 2 decades, GIC are increasing for certain subpopulations of CHD patients based on social determinants of health, including being older, lacking insurance, and residing in low Child Opportunity Index neighborhoods.

*What Are the Clinical Implications?:*

- GIC are worsening in certain subpopulations of CHD patients, including those with low Child Opportunity Index, lack of insurance, or poor access to health care systems.
- CHD programs must target social determinants of health factors to improve ongoing care and long-term outcomes for all patients.

## Introduction

Congenital heart disease (CHD) is the most prevalent birth defect in the United States, impacting around 0.8% of live births.^1^ As medical advancements have transformed complex CHD from a life-threatening condition to a chronic disease, the focus has shifted toward enhancing long-term outcomes through uninterrupted and comprehensive lifelong care. However, the persistent issue of gaps in care (GIC) among CHD patients remains, leading to clinical deterioration, hospitalizations, and increased mortality rates.^2–5^

Guidelines, including the American College of Cardiology, American Heart Association, European Society of Cardiology, and Canadian Cardiovascular Society, stress continuous care, especially during the pediatric to adult transition.^6–8^ Despite recommendations, GIC persist due to factors like perceived well-being, lack of symptoms, limited understanding of the necessity for follow-up, challenges accessing specialized care, and changes in insurance status.^9^ These underlying factors form a common thread that underscores the utmost importance of understanding social determinants of health (SDOH).^9^

Over the past decade, research has highlighted the critical role of SDOH in every stage of CHD, influencing early detection, overall incidence patterns, infant mortality, post-surgical recovery, and engagement with medical care, including follow-up appointments.^9–17^ While research over the past decade has highlighted the impact of SDOH at various stages of CHD, there is a noticeable gap in studies examining how GIC evolve and what factors are associated with these changes. Previous studies, while valuable, have limitations as they focus on specific SDOH factors and potentially miss the nuanced interactions between them.^9,11,13,17^

To address this gap, this study utilizes the Child Opportunity Index (COI), a comprehensive tool assessing education, health and environmental, and social and economic domains through 29 indicators.^18–20^ By utilizing COI and an extensive list of SDOH factors, this study aimed to not only understand the existing care gaps but also to explore the changing dynamics of these factors over time. This temporal analysis is crucial for gaining a more comprehensive understanding of the complex interplay of SDOH factors, how they change over time, and their potential impact on ongoing follow-up care for individuals with CHD.

## Methods

### Patient Selection

All CHD patients aged 0-34 years who underwent cardiac surgery at Nemours Children’s Health, Delaware Valley (NCHDV) in Wilmington, Delaware from January 2003 to May 2020 were included, utilizing data from the Society of Thoracic Surgeons (STS) database. The chosen start date aligns with NCHDV’s transition to electronic medical records, while the end date ensures at least 3 years of potential follow-up (at the time of analysis) for all patients for accurate evaluation of potential GIC. To identify continuity patients, we restricted our search to those with at least 1 cardiology follow-up appointment, aside from their surgical postoperative visit, and who resided within the region served by NCHDV (includes Delaware, Pennsylvania, New Jersey, and Maryland). To confirm that patients living in New Jersey and Maryland were our continuity patients, at least 5% of patients from these 2 states were reviewed in detail, revealing that 70% of those in New Jersey and 85% of those in Maryland were actively followed at NCHDV. Each patient’s cardiac diagnosis from the STS database was assigned a corresponding anatomic severity class: simple, moderate, or complex as defined by the 2018 American Heart Association/American College of Cardiology Guidelines for the Management of Adults with Congenital Heart Disease.^6^ For example, an isolated atrial septal defect was classified as simple, repaired tetralogy of Fallot or coarctation of the aorta as moderate, and single ventricle heart disease as complex (please refer to Supplemental Table 1 for additional examples). Aligned with the 2018 adult congenital heart disease (ACHD) guidelines, routine follow-up for most CHD patients is recommended within 2 years; for this analysis, we conservatively assumed this to be a reasonable approach for following pediatric CHD patients.

Consequently, irrespective of the specific CHD diagnosis, our cohort was anticipated to return for routine care within 3 years. A clinically significant GIC was defined as a lack of in-office or virtual follow-up, any visit related to further cardiac procedures or testing (eg, cardiac magnetic resonance imaging or cardiac catheterization), or patient phone calls within 3 years. This included both “observed gaps,” GIC between subsequent visits, and “ongoing gaps,” GIC that began at the patient’s most recent visit and were still ongoing as of May 20, 2023, the date on which the data were pulled. Deceased patients had the date of the data pull replaced with their date of death to ensure that the time after their death was not erroneously included in the GIC calculation.

### Data Collection

Institutional Review Board approval was completed prior to any data collection; by their guidance, informed consent was not required because of the retrospective nature of this study. The STS database was used to find cardiac surgery patients at NCHDV. Various SDOH factors were collected and analyzed as part of this study. Conversational fluency in English, ethnicity (Hispanic or non-Hispanic), and sex (male or female) were self-reported by patients and families at each of their office visits. The number of patient guardians was initially collected as a continuous variable before being converted into a binary format (at least 2 guardians or fewer than 2 guardians). Patient age was operationalized as a binary variable with a cutoff of 10.5 years; more details on the selection of this cutoff are given in the statistical analysis section. The patients’ primary race originally consisted of 17 levels; to facilitate the convergence of statistical models, these options were condensed and combined with ethnicity to produce the following 4 groups: non-White non-Hispanic, non-White Hispanic, White non-Hispanic, and White Hispanic.

We used the patient’s home address zip code in concert with the *zipcodeR* package^21^ to (1) determine the distance from the patient’s residence to the (i) nearest outpatient cardiology location and (ii) the tertiary care hospital in Wilmington, Delaware, and (2) to assign each patient a value for the COI.^18^ The patient’s reported address and zip code were compared with the COI 2.0 index data. The Institute for Children, Youth, and Family Policy at Brandeis University, Waltham, Massachusetts, created the COI database to measure 29 indicators of neighborhood conditions over 3 domains, including education, health and environmental, and social and economic, summed into 1 easily comparable score. These 29 indices are drawn from public sources such as the Census Bureau, the National Center for Education Statistics, the US Department of Agriculture, and the Environmental Protection Agency and look at factors such as household income, health insurance coverage, employment rate, and the experience level of teachers within the school district. The COI score ranges from 1–100 U and is categorized into very low (<20 U), low (20≤40 U), moderate (40≤60 U), high (60≤80 U), or very high (≥80 U) opportunity.^17–20,22^ Higher scores reflect more favorable neighborhood opportunities at the metropolitan, state, or national levels. COI scores were standardized at the metropolitan, state, and national levels. Details of the development of COI are available online.^22^ For our study, we divided COI into 3 main categories: very low/low (<40 U), moderate (40≤60 U), and high/very high (>60 U).

Patient insurance was first defined as public (Medicaid, Medicare, Managed Medicaid, and Tricare) or private (any other payor). From this, we created 2 additional variables: patient insurance composition (public only, private only, or mixed) and the number of unique payors over 1 year of the patient’s recorded medical history (0, 1, or 2+ years).

### Statistical Analyses

To create the analytic sample, we excluded any patients whose initial visits occurred after May 20, 2020 from the analysis, as it would not be possible for them to have experienced a GIC at the time of analysis. Further, patients receiving extracorporeal membrane oxygenation but lacking a cardiac diagnosis and patients who did not report living in 1 of the 4 states of interest (Delaware, Pennsyvlania, Maryland, and New Jersey) were also excluded. Approximately 20% of encounters had some missing information; patient-level missingness is summarized in Table 1.

**Table 1.**
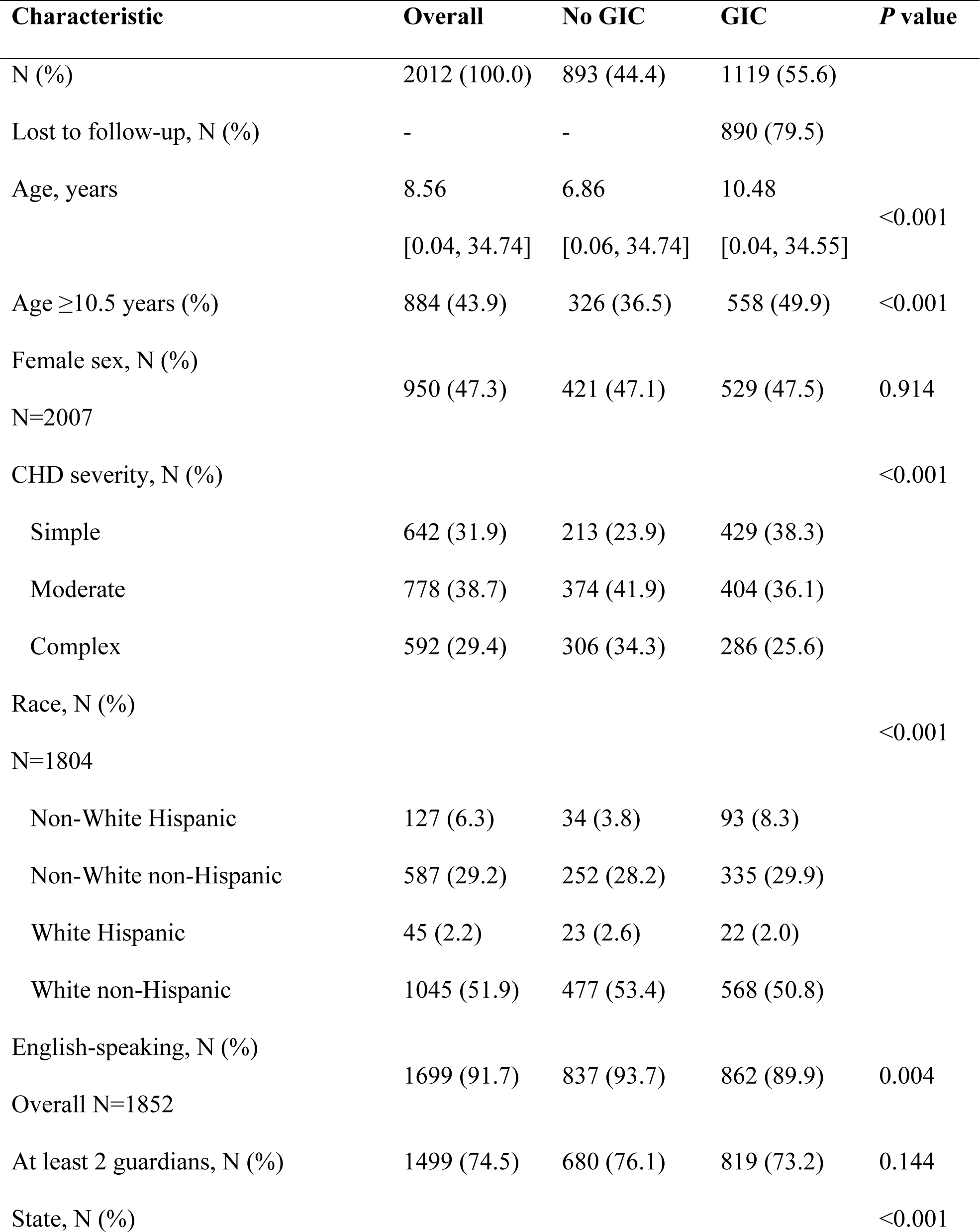
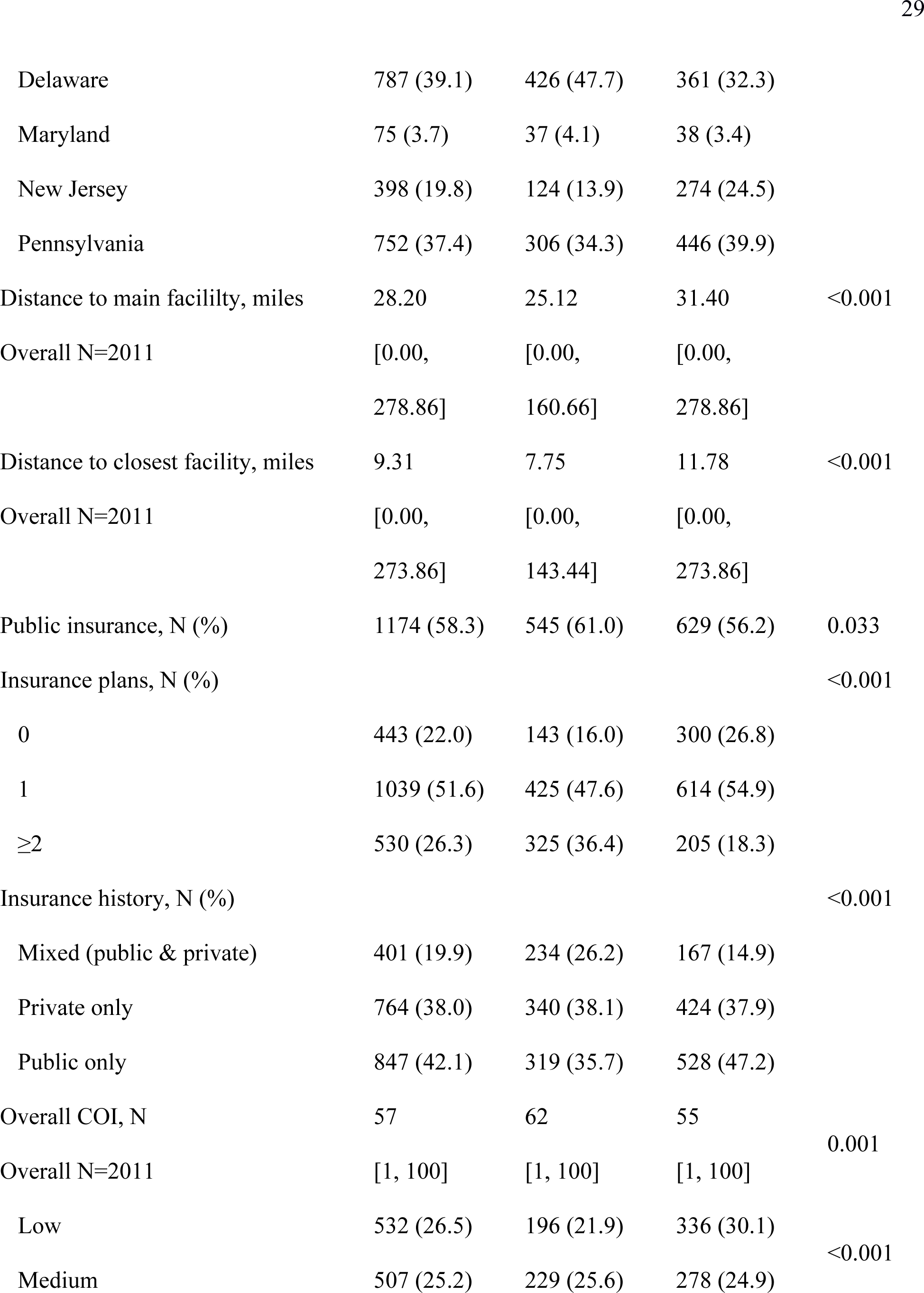
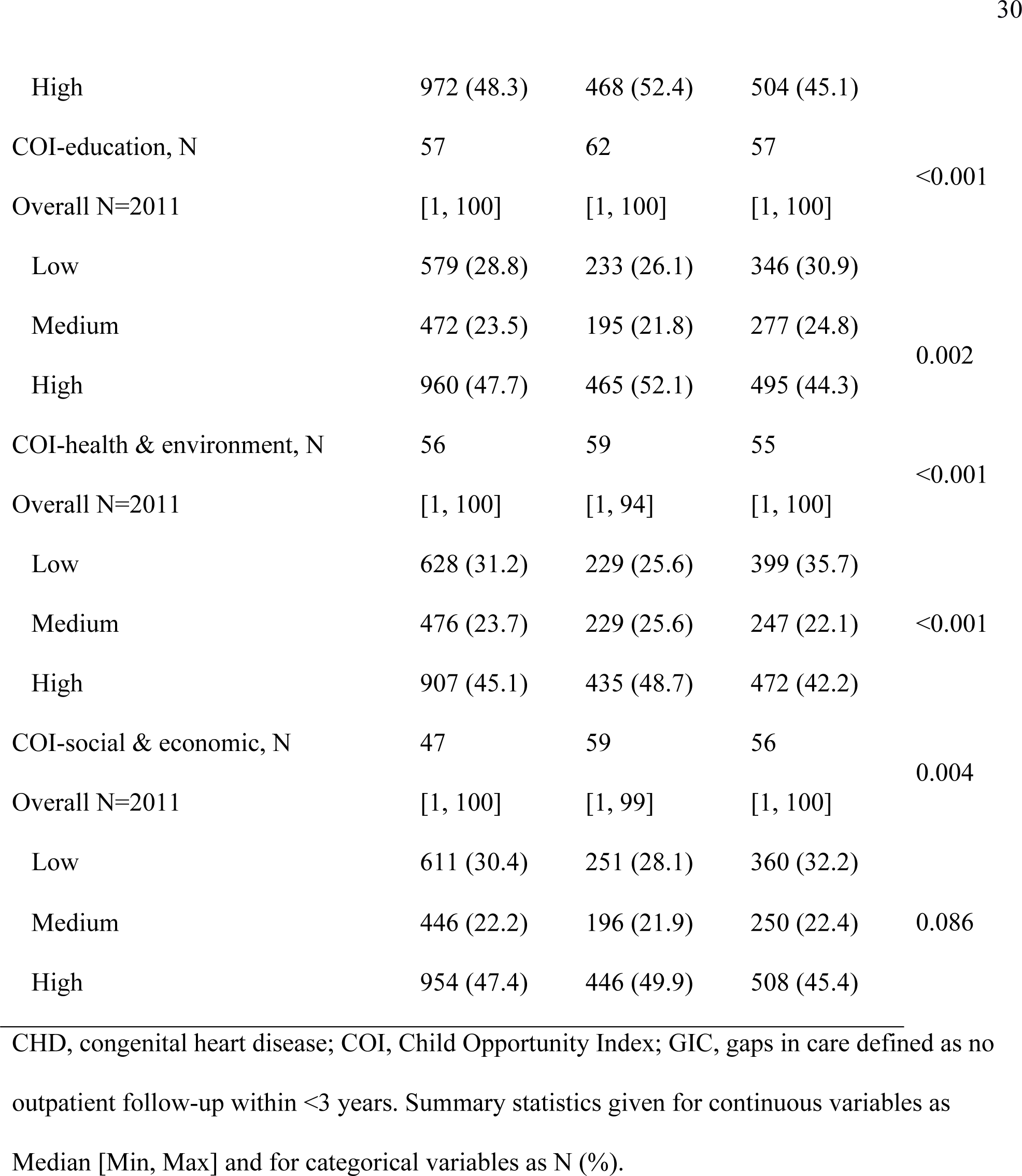
Demographic and Clinical Characteristics of CHD Patients with or Without GIC.

| **Characteristic** | **Overall** | **No GIC** | **GIC** | ***P* value** |
| --- | --- | --- | --- | --- |
| N (%) | 2012 (100.0) | 893 (44.4) | 1119 (55.6) |  |
| Lost to follow-up, N (%) | - | - | 890 (79.5) |  |
| Age, years | 8.56<br>[0.04, 34.74] | 6.86<br>[0.06, 34.74] | 10.48<br>[0.04, 34.55] | <0.001 |
| Age ≥10.5 years (%) | 884 (43.9) | 326 (36.5) | 558 (49.9) | <0.001 |
| Female sex, N (%) | 950 (47.3) | 421 (47.1) | 529 (47.5) | 0.914 |
| N=2007 |  |  |  |  |
| CHD severity, N (%) |  |  |  | <0.001 |
| Simple | 642 (31.9) | 213 (23.9) | 429 (38.3) |  |
| Moderate | 778 (38.7) | 374 (41.9) | 404 (36.1) |  |
| Complex | 592 (29.4) | 306 (34.3) | 286 (25.6) |  |
| Race, N (%) |  |  |  | <0.001 |
| N=1804 |  |  |  |  |
| Non-White Hispanic | 127 (6.3) | 34 (3.8) | 93 (8.3) |  |
| Non-White non-Hispanic | 587 (29.2) | 252 (28.2) | 335 (29.9) |  |
| White Hispanic | 45 (2.2) | 23 (2.6) | 22 (2.0) |  |
| White non-Hispanic | 1045 (51.9) | 477 (53.4) | 568 (50.8) |  |
| English-speaking, N (%) | 1699 (91.7) | 837 (93.7) | 862 (89.9) | 0.004 |
| Overall N=1852 |  |  |  |  |
| At least 2 guardians, N (%) | 1499 (74.5) | 680 (76.1) | 819 (73.2) | 0.144 |
| State, N (%) |  |  |  | <0.001 |
| Delaware | 787 (39.1) | 426 (47.7) | 361 (32.3) |  |
| Maryland | 75 (3.7) | 37 (4.1) | 38 (3.4) |  |
| New Jersey | 398 (19.8) | 124 (13.9) | 274 (24.5) |  |
| Pennsylvania | 752 (37.4) | 306 (34.3) | 446 (39.9) |  |
| Distance to main facility, miles | 28.20 | 25.12 | 31.40 | <0.001 |
| Overall N=2011 | [0.00, 278.86] | [0.00, 160.66] | [0.00, 278.86] |  |
| Distance to closest facility, miles | 9.31 | 7.75 | 11.78 | <0.001 |
| Overall N=2011 | [0.00, 273.86] | [0.00, 143.44] | [0.00, 273.86] |  |
| Public insurance, N (%) | 1174 (58.3) | 545 (61.0) | 629 (56.2) | 0.033 |
| Insurance plans, N (%) |  |  |  | <0.001 |
| 0 | 443 (22.0) | 143 (16.0) | 300 (26.8) |  |
| 1 | 1039 (51.6) | 425 (47.6) | 614 (54.9) |  |
| ≥2 | 530 (26.3) | 325 (36.4) | 205 (18.3) |  |
| Insurance history, N (%) |  |  |  | <0.001 |
| Mixed (public & private) | 401 (19.9) | 234 (26.2) | 167 (14.9) |  |
| Private only | 764 (38.0) | 340 (38.1) | 424 (37.9) |  |
| Public only | 847 (42.1) | 319 (35.7) | 528 (47.2) |  |
| Overall COI, N | 57 | 62 | 55 | 0.001 |
| Overall N=2011 | [1, 100] | [1, 100] | [1, 100] |  |
| Low | 532 (26.5) | 196 (21.9) | 336 (30.1) | <0.001 |
| Medium | 507 (25.2) | 229 (25.6) | 278 (24.9) |  |
| High | 972 (48.3) | 468 (52.4) | 504 (45.1) |  |
| COI-education, N | 57 | 62 | 57 |  |
| Overall N=2011 | [1, 100] | [1, 100] | [1, 100] | <0.001 |
| Low | 579 (28.8) | 233 (26.1) | 346 (30.9) |  |
| Medium | 472 (23.5) | 195 (21.8) | 277 (24.8) |  |
| High | 960 (47.7) | 465 (52.1) | 495 (44.3) | 0.002 |
| COI-health & environment, N | 56 | 59 | 55 |  |
| Overall N=2011 | [1, 100] | [1, 94] | [1, 100] | <0.001 |
| Low | 628 (31.2) | 229 (25.6) | 399 (35.7) |  |
| Medium | 476 (23.7) | 229 (25.6) | 247 (22.1) | <0.001 |
| High | 907 (45.1) | 435 (48.7) | 472 (42.2) |  |
| COI-social & economic, N | 47 | 59 | 56 |  |
| Overall N=2011 | [1, 100] | [1, 99] | [1, 100] | 0.004 |
| Low | 611 (30.4) | 251 (28.1) | 360 (32.2) |  |
| Medium | 446 (22.2) | 196 (21.9) | 250 (22.4) | 0.086 |
| High | 954 (47.4) | 446 (49.9) | 508 (45.4) |  |

CHD, congenital heart disease; COI, Child Opportunity Index; GIC, gaps in care defined as no outpatient follow-up within <3 years. Summary statistics given for continuous variables as Median [Min, Max] and for categorical variables as N (%).

Summary statistics were generated for all variables of interest to characterize the sample: means and standard deviations for continuous variables and frequencies and percentages for categorical variables. These data were also stratified by overall GIC status (none or at least 1) to determine if the composition of the study variables differed. Any statistically significant differences were identified using t-tests for continuous variables and chi-square tests for categorical variables.

Before fitting any models, the minimized Euclidean distance was employed with a receiver operating characteristic curve to determine the optimal age cutoff in the GIC prediction. Age was then parameterized as a binary variable, with the 2 groups consisting of (1) individuals the same age or younger than the optimal cutoff and (2) individuals older than the optimal cutoff. To assess the nature of the relationship between GIC and time, CHD severity, and the SDOH factors of interest, we fit 2 generalized linear models utilizing generalized estimating equations, 1 featuring the entire analytic sample and a second featuring a subsample of patients with moderate and complex CHD only. Given the binary nature of the outcome variable, the logit link function was used. An autoregressive working correlation structure was assumed in anticipation of weakening similarity between successive measurements made on a given patient over time.

Fortunately, the beta coefficients estimated using generalized estimating equations possess many desirable statistical properties, even in the case of the working correlation matrix’s misspecification; therefore, this choice only needs to reflect the expected correlation structure roughly.^23^ This model excludes missing encounters from analysis; however, other nonmissing encounters from the same patients are retained. Model results were reported using odds ratios, corresponding 95% confidence intervals, and *P* values. Data cleaning and summary statistic generation were performed in R version 4.3.1.^24^ Multivariable models were fit using SAS (Cary, North Carolina) version 9.4. The level of significance was set at 0.05.

## Results

### Subjects

Two thousand and twelve patients with surgically repaired CHD who were seen between 2003 and 2020 and resided within Delaware, Pennsylvania, New Jersey, and Maryland formed our cohort of continuity patients (Table 1). Of the overall cohort, the median age was 8.56 years, with 47.3% female; 51.9% were White non-Hispanic. Most patients resided in Delaware (39.1%) and Pennsyvlania (37.4%); the remaining patients lived in either New Jersey or Maryland (23.5%). Of this group, CHD diagnosis level was complex in 29.4%, moderate in 38.7%, and simple in 31.9%. Most patients (58.3%) were on public insurance, with 26.3% reporting multiple insurance plans.

### Comparison of Patients with and Without a GIC

Of the total cohort, 1119 (55.6%) had a GIC greater than 3 years at any given point in their ongoing care (Table 1). Univariate analysis showed CHD patients with GIC were more likely to be older and live outside the state of Delaware, specifically in New Jersey or Pennsylvania.

Those with GIC were more likely to have simple CHD; however, even among those with moderate and complex CHD (1370 patients), ∼50% had a GIC. Patients with a GIC were more likely to be non-English speaking, non-White, and of Hispanic ethnicity. In terms of insurance, patients with a GIC were most likely to be on public insurance only and least likely to be on both public and private insurance. Those without GIC tended to have more insurance coverage, with 36.4% having 2 or more insurance policies over their entire medical history. Among those with GIC, only 18.3% had 2 or more insurance policies before their GIC.

### GIC Longitudinal Trends

Through a comprehensive longitudinal analysis, we observed a general increase in GIC for all CHD patients, regardless of complexity (Figure 1). Notably, the highest proportional rise was evident in patients with simple CHD (average 1.1% per year). Utilizing our 2 longitudinal multivariable models (Tables 2 and 3) and dissecting GIC trends among subgroups with specific risk factors such as age, insurance, access, and COI, we discovered that these factors primarily drive the escalating GIC trend. Specifically, in subpopulations with these risk factors, GIC is on the rise, while patients lacking these risk factors show an overall significant decrease in GIC over time (Figure 2 and Supplemental Figure 1).

**Figure 1.**
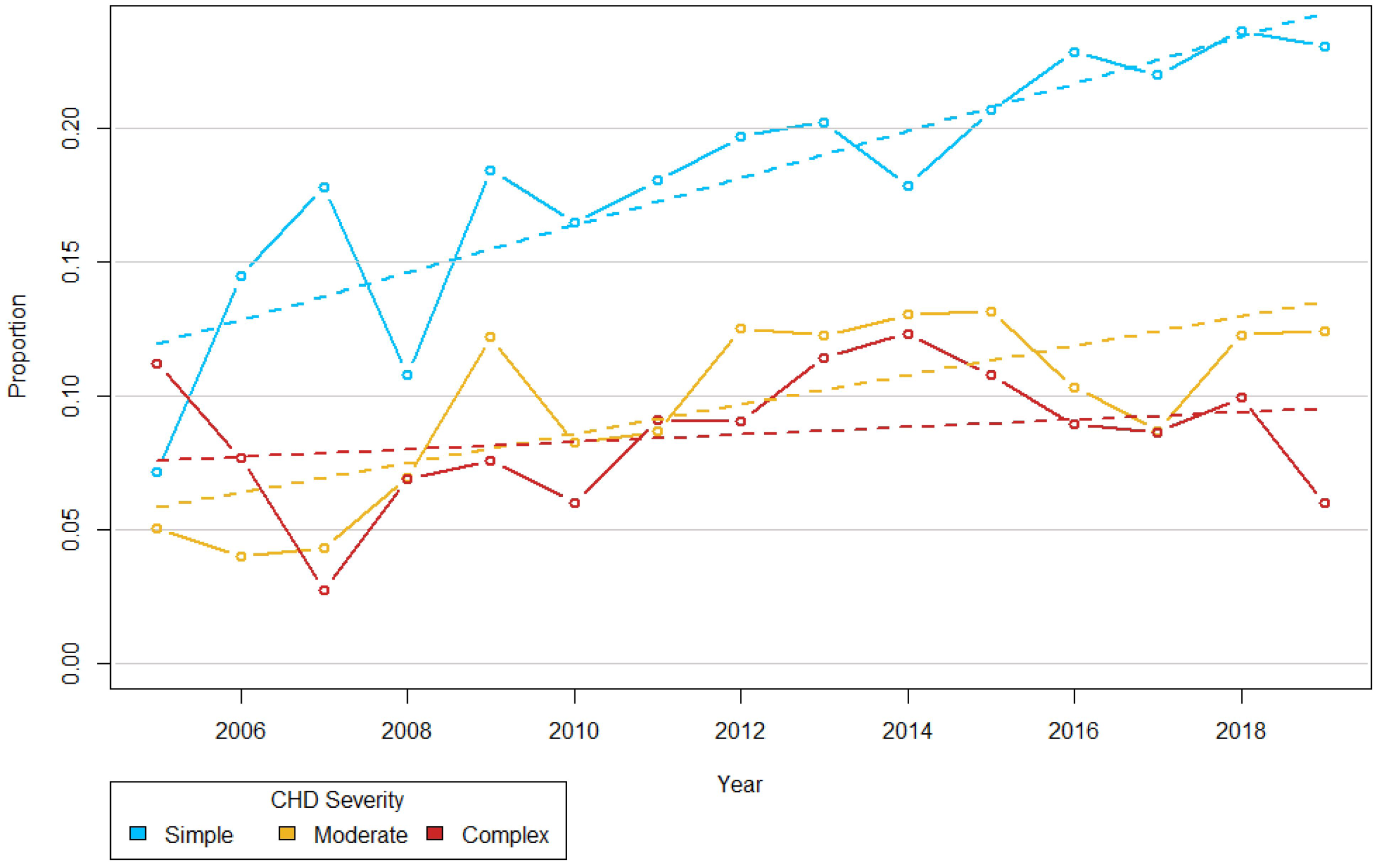
Graph showing the increase in proportion of patients with congenital heart disease with gaps in care every year for patients with simple (blue line), moderate (orange line), and complex (red line) congenital heart disease.

**Figure 2.**
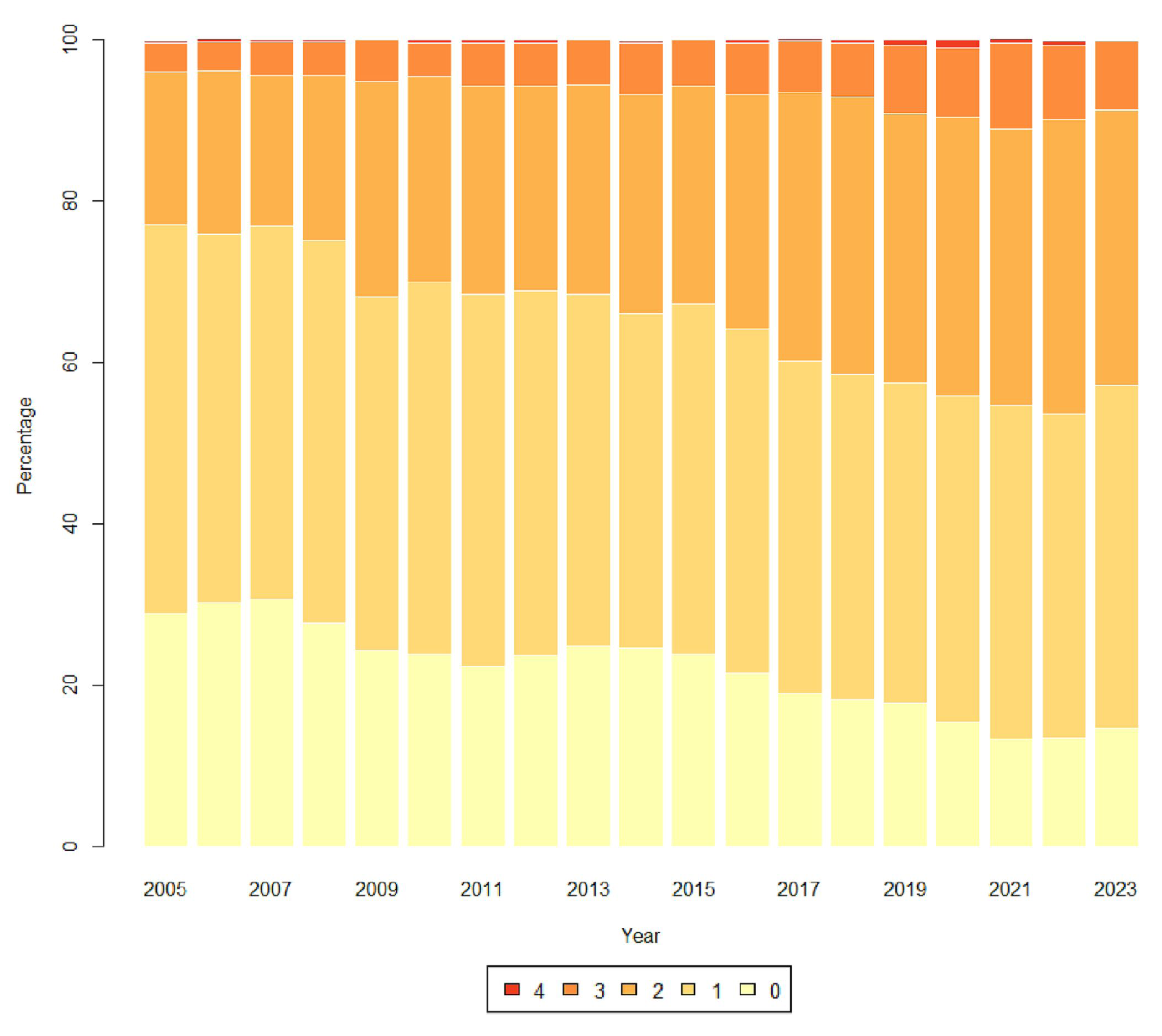
Graph showing an increase in the number of patients with congenital heart disease and the distribution and number of social determinants of health risk factors over time. The risk factors are (1) age ≥10.5 years, (2) living in New Jersey or Pennsylvania, (3) simple congenital heart disease, (4) no insurance plans, and (5) low health and environment Child Opportunity Index group.

**Table 2.**
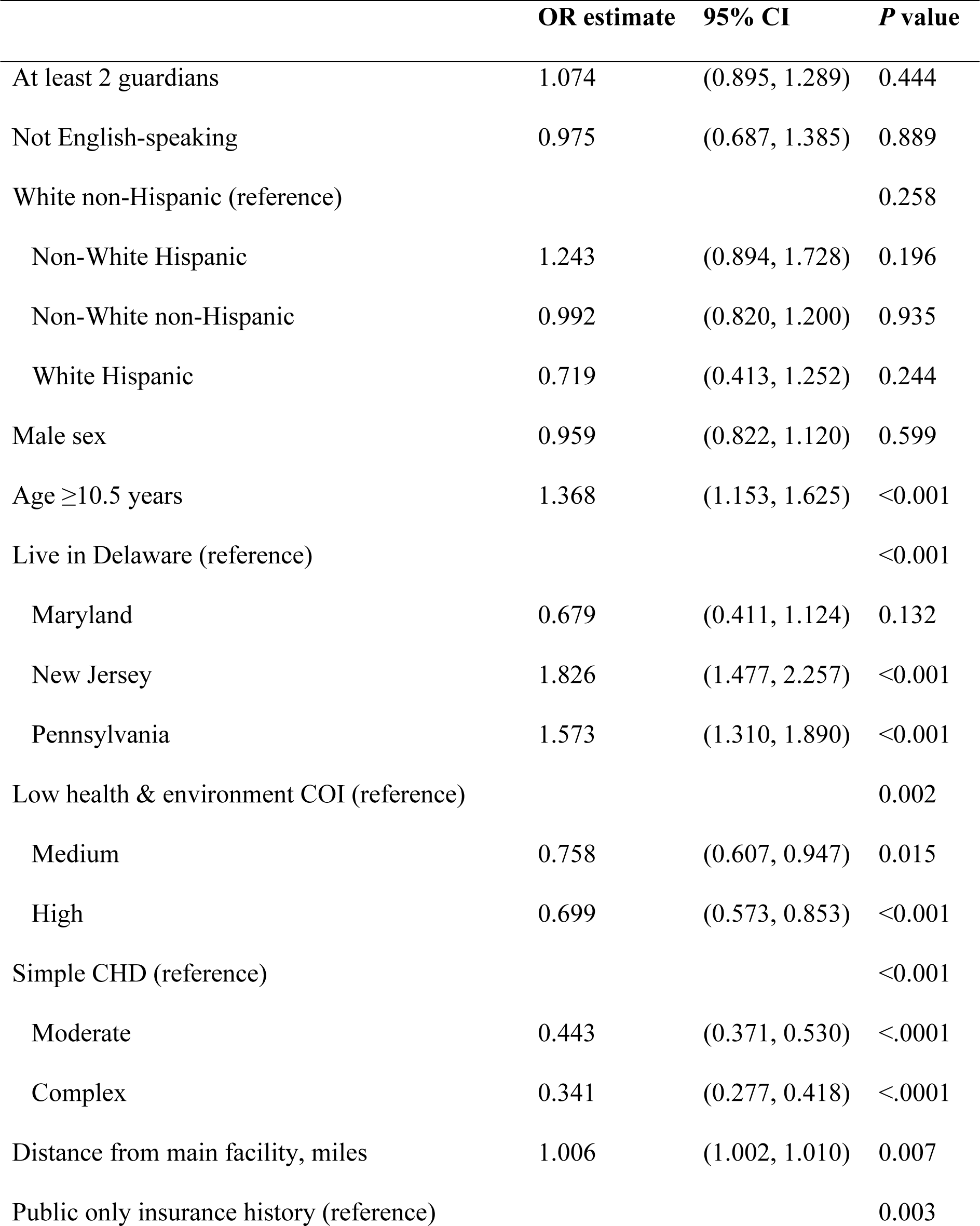
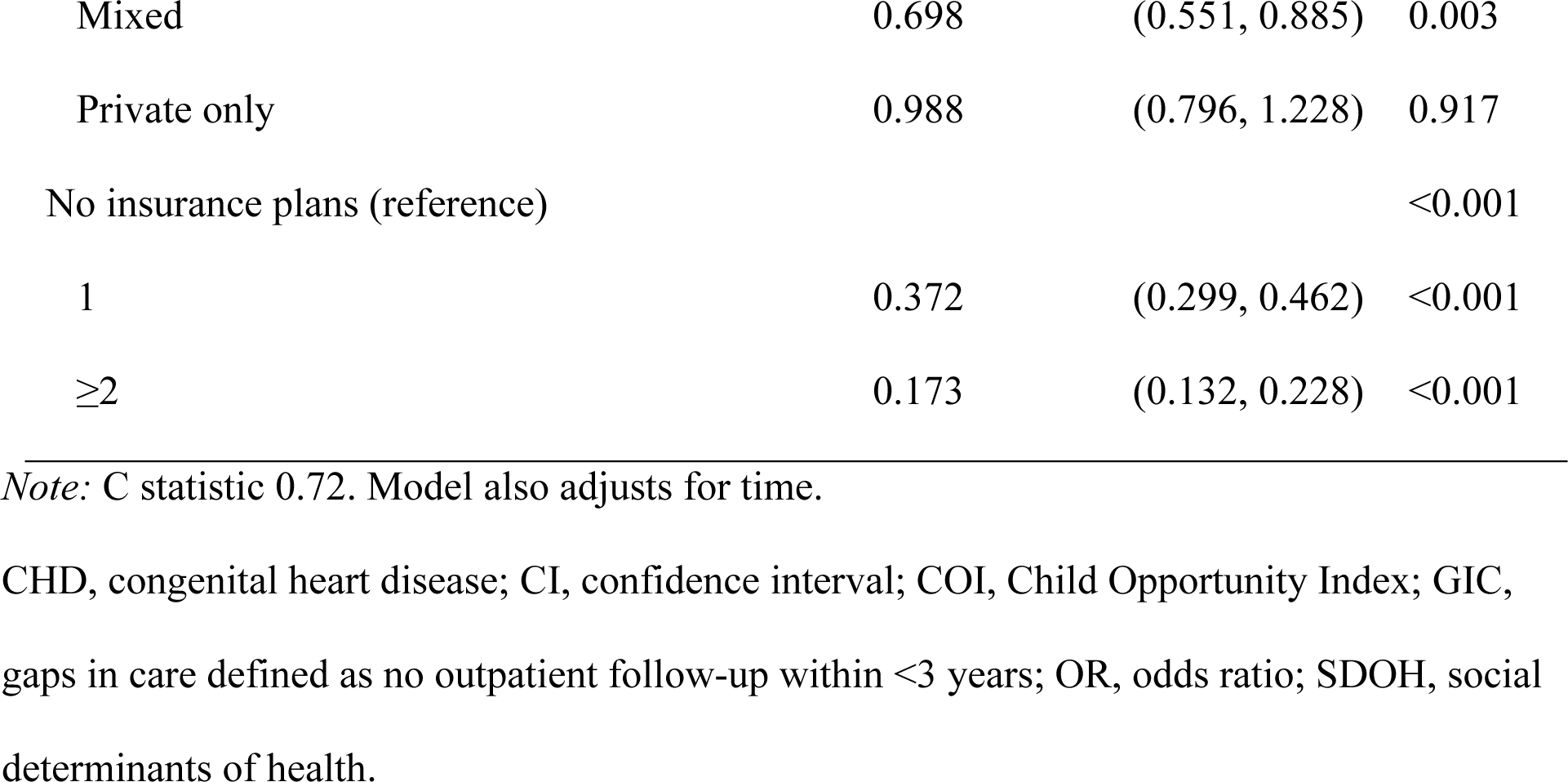
Multivariable Longitudinal Model Assessing SDOH Factors Associated with GIC Over Time.

**Table 3.** Multivariable Longitudinal Model Assessing SDOH Factors Associated with GIC Over Time for Moderate and Greatly Complex CHD Patients Only.

|  | **OR estimate** | **95% CI** | **P value** |
| --- | --- | --- | --- |
| At least 2 guardians | 1.105 | (0.872, 1.401) | 0.409 |
| Not English-speaking | 0.982 | (0.614, 1.568) | 0.938 |
| White non-Hispanic (reference) |  |  | 0.150 |
| Non-White Hispanic | 1.418 | (0.952, 2.111) | 0.086 |
| Non-White non-Hispanic | 0.919 | (0.717, 1.176) | 0.500 |
| White Hispanic | 0.630 | (0.252, 1.575) | 0.323 |
| Male sex | 1.016 | (0.830, 1.243) | 0.881 |
| Age $\geq 10.5$ years | 1.425 | (1.153, 1.761) | 0.001 |
| Live in Delaware (reference) |  |  | <0.001 |
| Maryland | 0.790 | (0.431, 1.447) | 0.445 |
| New Jersey | 1.865 | (1.420, 2.450) | <0.001 |
| Pennsylvania | 1.508 | (1.185, 1.918) | 0.001 |
| Low health & environment COI (reference) |  |  | <0.001 |
| Medium | 0.689 | (0.521, 0.912) | 0.009 |
| High | 0.586 | (0.458, 0.749) | <0.001 |
| Complex CHD severity | 0.734 | (0.596, 0.904) | 0.004 |
| Distance from main facility, miles | 1.010 | (1.007, 1.013) | <0.001 |
| Public only insurance history (reference) |  |  | 0.007 |
| Mixed | 0.639 | (0.467, 0.872) | 0.005 |
| Private only | 0.966 | (0.730, 1.277) | 0.807 |
| No insurance plans (reference) |  |  | <0.001 |
| 1 | 0.399 | (0.301, 0.529) | <0.001 |
| $\geq 2$ | 0.171 | (0.120, 0.245) | <0.001 |

*Note:* C statistic 0.72. Model also adjusts for time.
CHD, congenital heart disease; CI, confidence interval; COI, Child Opportunity Index; GIC, gaps in care defined as no outpatient follow-up within <3 years; OR, odds ratio; SDOH, social determinants of health.

### COI and interactions with GIC

The overall median COI for the entire cohort was 57. On univariate analysis, COI was significantly lower in patients with GIC overall and all subdomains. When COI was divided into subcategories of low, moderate, and high, patients with a GIC were more likely to have a low COI compared with those with no GIC.

### Longitudinal model of all CHD patients with GIC

Examining rates of GIC per year, being older, having simple CHD, living outside of Delaware, living farther from the main hospital, being on public insurance and with no insurance plans, and living in low COI neighborhoods for the health and environment subdomain were associated with increasing rates of GIC over time, independent of other factors in the more recent era (Table 2).

### Longitudinal model for moderate and complex CHD patients with GIC

The same findings were seen when patients with simple CHD were excluded; longitudinal modeling showed rates of GIC increased over time, associated with older age, living outside of Delaware, living farther from the main hospital, residing in low COI neighborhoods for the health and environment subdomain, and being on public insurance and fewer insurance plans (Table 3).

## Discussion

This retrospective analysis, spanning 2 decades and involving 2012 patients with repaired CHD, reveals that over half experienced GIC of 3 or more years. When examining GIC over time, all types of CHD diagnoses had an increasing GIC over time. Proportionally, those with complex anatomy had few GIC in contrast to those with moderate and simple anatomy, who displayed higher proportions of GIC. On further analysis, increasing GIC trends were also identified in older patients residing outside Delaware, farther from the main hospital, having public or limited insurance coverage, and living in neighborhoods with low COI.

Previous studies have provided valuable insights into the impact of SDOH factors on CHD outcomes, including GIC, but there are certain limitations to consider. The studies concentrated on specific factors including poverty, low socioeconomic status, parental education, and transportation barriers; other potential SDOH influences are likely at play.^9,11,13,17^ Additionally, while the studies highlight associations, they may not fully capture the intricate complexity of these relationships, as certain factors might interact in nuanced ways. Given the dynamic nature of certain SDOH factors, they may change over time.

Our study aligns with existing literature by affirming the correlation between advancing age and insurance challenges with contributing to higher rates of GIC.^25^ However, our study adds unique insights into GIC, highlighting the persistent and escalating trend, independent of CHD severity, in patients with specific SDOH characteristics. This underscores the intricate interplay between broader societal dynamics and a health care system potentially ill-equipped to address these unattended issues, which likely contributes to the observed rise in GIC within this patient subgroup. The findings emphasize the ongoing challenge of GIC, particularly in specific CHD subpopulations lacking adequate mitigation measures.

Despite advances in clinical management and a substantial decrease in mortality, CHD patients continue to face significant morbidity and poor long-term outcomes associated with GIC.^26^ Recognized as an independent predictor of mortality, GIC are 3 times more likely to lead to urgent interventions, patients receiving a new cardiac diagnosis, and patients presenting symptomatically.^26^ Our analysis reveals that GIC in certain subpopulations are ongoing and increasing. In the absence of interventions that proactively incorporate or focus specifically on SDOH factors in susceptible subpopulations of CHD patients, the trajectory of these unfavorable outcomes is unlikely to be reversed.

Our data showed that GIC are more common after the age of 10.5 years, which is congruent with previously published literature suggesting that GIC in those with CHD commonly begin in early adolescence and increase in length and occurrence into the early 20s, with almost half of those with CHD having a significant GIC by the age of 20 years.^27^ Frequent reasons cited are lack of access to ACHD programs, lapses in insurance, a lack of transition education, and a lack of transfer of care.^5,26,28–32^ Males are commonly thought to be more likely to experience a GIC; however, our data and previous studies have not shown significant differences based on sex.^27,33^ Distance to the site of care has been shown to correlate with GIC.^28^ NCHDV’s health care system has 1 primary facility with 9 satellite campuses spanning 3 states. Within this population, distance from NCHDV and its satellite campuses was associated with GIC.

Nonmodifiable SDOH, including attributes like race and ethnicity, have shown a connection to GIC.^27^ However, it is crucial to acknowledge that these factors only partially capture the intricate web of societal inequalities, systemic biases, and disparities. Our multivariable model revealed a significant shift where the importance of race and ethnicity diminished independently of other risk factors. Instead, factors such as COI and insurance, indicative of broader societal disadvantages, emerged as pivotal contributors. This suggests that although race and ethnicity play a role, they are imperfect surrogates for social inequities explaining GIC. The interplay of complex factors necessitates a holistic understanding beyond surface-level demographics.

Additionally, certain racial groups, such as Asian Americans and Pacific Islanders, were represented by relatively small overall populations. The limited sample size warrants caution when drawing conclusions about specific racial groups within the context of the study’s findings.

Issues related to insurance and health care access are acknowledged contributors to heightened GIC for patients with CHD.^28^ Our study additionally reveals that relying solely on public insurance, without supplementing additional insurance or a co-insurance plan, is independently linked to an increased prevalence of GIC. This finding suggests that even within the subgroup of patients covered by public insurance, disparities in resources may exist, such as having multiple insurances through employment or familial support, providing added financial protection.

Additionally, certain families may have sought support from social work services, enhancing their ability to secure and navigate these essential insurance plans. Therefore, it is essential to understand the details of the variability of access to social work interventions and how they may have impacted GIC for certain families.

Although previous studies indicated lower COI scores were associated with worse outcomes for CHD patients,^19^ our study brings a novel perspective. Specifically, within the COI, we observed that the subdomain related to health and environment independently correlates with GIC. A notable indicator within this subdomain is the percentage of health insurance coverage in the population, emphasizing the profound connection between this aspect of the COI and GIC. This underscores the crucial role of health care access, suggesting that other subdomains associated with socioeconomic factors (eg, poverty rate, single-parent households, and employment) or education (such as the number of high school graduates in the neighborhood) may have a diminished impact on ongoing follow-up. Instead, insurance and health care access emerge as pivotal determinants in GIC trends.

Moreover, the aging ACHD population introduces a unique dynamic where CHD patients undergo insurance transitions as they age.^34^ This transition is a critical factor that deserves careful consideration during ACHD counseling and the transition of care. Addressing insurance-related challenges becomes increasingly essential as CHD patients age, ensuring seamless access to health care services and minimizing disruptions in follow-up care.

Unlike the ACHD population, there are no formal guidelines for pediatric CHD follow-up. However, guidelines for managing adults with CHD recommend that the pediatric population be followed routinely (every 2 years), and transition education including insurance, medical history, and directed transfer of care be provided to help reduce GIC and loss of follow-up.^6,35^

The lack of formal pediatric CHD follow-up guidelines and anatomical and clinical variability has resulted in significant practice variation, where providers often recommend follow-up intervals based on individual practices. There is a tendency to perceive patients who underwent CHD surgery, especially those with simple or moderate but now repaired CHD, as “fixed.” However, the prevalence of adults living with CHD surpasses that of children, and these perceptions contribute to ongoing GIC. Studies have shown that even simple CHD is not benign, with patients at risk for significant morbidity and mortality compared with controls.^36^ Patients with simple CHD are at lifelong risk for worsening anatomical lesions needing surgical or cath-based interventions, arrhythmias, heart failure, and sudden death.^28,36^

### Implications

Despite advances in CHD care, persistent gaps contribute to adverse outcomes. An analysis spanning 2 decades reveals worsening GIC in specific subpopulations, indicating a potential for poorer outcomes. The growing proportion of adults with CHD faces challenges like inadequate health care access, highlighting socially and economically complex patient trends. As the aging population presents intricate SDOH needs, there is a compelling need to reassess CHD care delivery. Social inequities may lead to varying levels of ongoing care, emphasizing the critical need for tailored care strategies to improve care for all CHD patients despite societal disadvantages.

Preventing GIC may involve proactive symptom investigation, ongoing provider education, and frequent follow-up regardless of the severity of CHD. Screening and addressing high-risk SDOH factors are essential corroboration with cardiac management and may require frequent clinic visits. Transition programs focusing on SDOH factors, building patient independence from aging patients, helping patients navigate social challenges, and focusing on preventative health measures are crucial for addressing nonanatomical cardiac risk factors.

Efforts to decrease GIC for CHD patients will require a multifaceted approach, with social workers, care coordinators, and nurse educators crucial in providing uninterrupted lifelong care by using, eg, a learning health system approach.^37^ Additionally, there is a need for a deeper understanding of electronic medical records to screen for risk factors associated with GIC, potentially leveraging artificial intelligence and natural language processing modalities. In the absence of a proactive incorporation of SDOH factors, particularly for susceptible subpopulations among CHD patients, the persistence of GIC and poor outcomes seems inevitable. It is no longer just an option but an imperative to address these disparities directly. A comprehensive approach that integrates SDOH factors is vital for achieving better outcomes and fostering more equitable health care for CHD patients, especially those navigating the intersection of vulnerability and inadequate care.

## Limitations

Our study possesses several inherent limitations. It relies on a retrospective dataset from a single tertiary center study, utilizing information collected in routine clinical care. The sociodemographic data, including race, ethnicity, and preferred language, were self-reported during routine office visits, allowing patients or family members to opt out of specific questions. These limitations are even more evident, particularly in the nuances of racial and ethnic identities being captured in large, broad categories. For instance, the term “Asian” lacks specificity and overlooks the distinction between multiple cultures housed within the group, such as the Japanese, Chinese, and Indian cultures, each of whom has a unique perspective on medical culture. Recognizing and addressing these nuances is crucial in obtaining culturally sensitive demographic information. The accuracy of medical records, particularly regarding language preferences or translator usage, may vary as the data were not collected for research purposes.

The categories pertaining to race and ethnicity mirror those found in the US Census, which carries inherent limitations due to outdated definitions that fail to adequately capture the diversity of the US population. Due to the large number of patients, individualized follow-up recommendations for patients were not considered, potentially leading to variations with some patients possibly advised for a follow-up period exceeding 3 years and others advised to return earlier. Our analysis does not factor in patient outcomes, such as death or relocation, following their last routine office visit. There is a possibility that GIC observed in the data could be attributed to patients who have passed away or relocated, but this information remains unknown in our database. Furthermore, explanations for GIC may include patients moving to a different region or a transfer of care, and we did not reach out to patients with GIC or their primary care providers for additional insights. In contrast, the dataset lacked representation of those who entered the health care system, did not need surgical intervention, or intervened at an outside facility. This omission raises the need for further consideration and evaluation of those who enter health care systems later in life or after corrective repairs. Lastly, the COI was restricted to components for which nationally representative data were available, limiting the scope of this index in capturing a comprehensive picture of SDOH in our study population.

## Data Sharing Statement

All data, analytic methods, and study materials will be available to other researchers for data analysis to reproduce the results or replicate the procedure upon request. This data will be deidentified, and no personal health information will be shared.

## Sources of Funding

None.

## Disclosures

None.

## Acknowledgements

Author contributions: All authors critically reviewed and contributed to the intellectual property of the manuscript. Abbas H. Zaidi, Claude Beaty, Jr., Devyani Chowdhury, Sarah D. de Ferranti were involved with the concept of this study. Initial data preparation was done by Adam Alberts, Benjamin Brewer, with statistical analysis being done by Benjamin Brewer. Abbas H. Zaidi, Adam Alberts, Benjamin Brewer drafted several variations of the manuscript. Devyani Chowdhury, Claude Beaty, Jr., Sarah D. de Ferranti, Ming Hui Chen, Abbas H. Zaidi, MD provided clinical expertise and edited the manuscript for important intellectual content.

## Nonstandard Abbreviations and Acronyms

ACHD: Adult congenital heart disease
CHD: Congenital heart disease
COI: Child Opportunity Index
GIC: Gaps in care
NCHDV: Nemours Children’s Health, Delaware Valley
SDOH: Social determinants of health
STS: Society of Thoracic Surgeons

## Notes

### Competing Interest Statement

The authors have declared no competing interest.

### Funding Statement

No external funding was received in support of this work.

### Author Declarations

This study was approved by the Nemours Institutional Review Board.

